# Public attitudes towards protecting Emergency Medical Services staff from violence and aggression: a qualitative content analysis of free-text survey responses

**DOI:** 10.64898/2026.11.25.26368297

**Authors:** Nigel Rees, Daniel Todd, Claire Hawkes, Lauren Williams, Julia Williams, Peter O’Meara

## Abstract

**Background:** Violence and aggression (V&A) occurs in approximate 4-5% of Emergency Medical Services (EMS) calls. EMS staff endure many forms of V&A including verbal threats, physical injury, sexual assaults and on rare occasions fatalities occur. This can result in stress, fear, anxiety, and burnout in EMS staff, and there is significant interest in tacking this issue. Few evidence-based interventions exist, and despite their importance, public attitudes are notably missing in this discourse. This paper presents findings of a qualitative content analysis of free text survey responses to a survey of Wales (UK) adult attitudes towards V&A directed towards EMS staff.

**Methods:** Two independent samples of online YouGov surveys of Wales adults conducted in 2022 and 2024 were combined and qualitative content analysis conducted of the free text narrative responses.

**Results:** Two thousand and fourteen narrative responses were analysed, and the following conceptual themes emerged:

- *Multimodal, cumulative and confabulated knowing*
- *Dual discourse of V&A directed towards EMS staff*

**Conclusion:** The public overwhelmingly reported disgust at V&A towards EMS staff and appear to recognise the complexity of this issue. Whilst responses reflect efforts aimed at tackling this issue, including media campaigns and legislation such as the Assaults on Emergency Workers (Offences) Act (2018), we found limited awareness of the actual legislation and media campaigns. Rather, public attitudes appear to be influenced through multimodal and cumulative processes including social interaction, personal views and reflection.

We found support for prosecution and harsh criminal sentences for perpetrators of V&A towards EMS staff which aligns to retributivist perspectives and suggests public policy may be aligned with societal and moral expectation. We found limited evidence however for the preventative capacity of current approaches, which is consistent with our previous findings and wider body of knowledge. We therefore call for future research and policy to focus on the complexity and dual discourse revealed in this and other studies and follow consequentialist approaches to tackling these issues to prevent future V&A, reduce recidivism, and promote safety.

## Introduction

Violence and aggression (V&A) are one of the most common threats Emergency Medical Services (EMS) staff encounter within the workplace. Up to 90% of EMS staff report experiencing some form of V&A during their work and this is also present in 4-5% of 999 calls (Bigham et al 2014, Grange & Corbett 2002, Boyle et al 2007, Murray et al 2019). EMS staff experience many forms of V&A, including verbal threats, physical injury (sometimes involving weapons), sexual assaults and on rare occasions fatalities occur (Maguire et al 2018). Exposure to such V&A can result in increased levels of stress, fear, anxiety, emotional exhaustion, and burnout (Taylor et al 2016, Yoon et al 2016, Bernaldo-De-Quiros et al 2015).

EMS staff have a complex relationship with society, protecting public health while also being exposed to personal and psychological harms including V&A. This presents challenges in balancing complex clinical, societal and ethical issues while maintaining the reciprocal duty to protect EMS staff. Maguire (2018) contends that effective solutions to the growing problem of V&A directed toward staff are multifaceted, and include training, community education, adjustments to policies and national commissions are needed.

The PEACE collaborators include a range of international EMS clinicians, leaders and academics who have developed a program of research and innovation which aims to explore protecting EMS staff from aggression and violence in conflict encounters. We have described how this issue has become a priority in Wales (UK) where the multi-agency approach adopted has involved the UK and Welsh Governments, the Welsh Ambulance Services University NHS Trust (WAST), Health Boards, the NHS Wales Anti-Violence Collaborative, and the Joint Emergency Services Group in Wales (Rees et al 2021). We have also described how policy developments and initiatives from these groups include changes in legislation such as the Assaults on Emergency Workers (Offences) Act (2018), the Obligatory Responses to Violence in Healthcare (2018), and the communication campaign #WithUsNotAgainst Us (WAST 2026). The internationally published body of knowledge is however limited, few evidence-based interventions exist, and despite their importance, public attitudes to this issue are notably missing. We recently published the quantitative results of a survey that aimed to explore public attitudes toward Protecting Emergency Medical Services (EMS) Staff from Aggression and Violence in Conflict Encounters (Rees et al 2025). This paper presents a qualitative content analysis of the free text survey responses.

## Methods

An online YouGov survey was conducted between 18th - 24th August 2022 and then again between 1^st^ - 5^th^ February 2024. The surveys included 2014 Wales adults (aged 18+) [2022 n=1,010 and 2024 n=1004] from a matched panel representative of the population derived from a UK YouGov panel of >360,000 adults registered and separately weighted to be representative of adults in Wales in terms of social class, age, sex, and type of newspaper chosen (YouGov 2022). Researchers [CH, NR] familiar with this method (Hawkes et al 2017) developed the survey and worked with YouGov to optimize question clarity of meaning and ease of understanding. Survey questions were developed from a literature review, published in previous work (Rees et al 2021), and correspondence and discussion with people and groups with expertise in this area.

We have previously published the quantitative results from the 2022 survey (Rees et al 2025) and in this paper we combine independent samples of these surveys [2022 n=1,010 and 2024 n=1004] and report qualitative content analysis of the free text narrative responses to the following questions within both the 2022 and 2024 surveys:

- You previously told us that violence and/or aggression towards EMS staff can be acceptable in some cases… What are the reasons why you think this?
- You previously said that you have heard about violence and/or aggression directed towards EMS staff… In which, if any, of the following situations have you ever heard about violent and/or aggressive incident/s?
- You previously told us that you were aware of recent publicity and/or media campaigns concerning respect towards emergency workers… What was the main focus of the campaign(s)? (Please type your answer(s) in the box below, giving as much detail as possible)

Respondents could answer in the Welsh language and are translated I brackets in text.

### Analysis

Data were analysed using conventional content analysis (Hsieh and Shannon (2005). With limited existing knowledge on the topic, this method’s inductive approach was appropriate where the data is coding and then grouped into categories and themes. Analysis followed a systematic process of familiarisation, coding, categorisation, thematic analysis and interpretation. Survey results were read repeatedly to become familiar with the content and to gain an overall understanding of respondents’ responses. Textual responses to each question’s data were analysed separately and entered onto a coding book. Codes were compared to identify similarities, differences, and recurring patterns and assigned codes that captured their meaning and relevance to the research question. Related codes were subsequently grouped into broader categories and, where meaningful relationships were identified, developed into overarching themes.

Throughout the analysis, attention was given to both the explicit content of respondents’ accounts and the context in which statements were made. Coding was reviewed iteratively to ensure that categories remained grounded in the original data and accurately represented respondents’ perspectives. The researcher also revisited the surveys during the analysis to check that interpretations were supported by the data and that relevant or contradictory accounts were not overlooked. This approach provided a structured yet flexible framework for identifying patterns within the dataset while retaining the context and meaning of respondents’ responses.

Trustworthiness was observed through reflexivity, maintaining memos and an audit trail documenting assumptions and monitoring any biases that may affect interpretation and each step of the coding decisions. A coding book was developed, and a second researcher reviewed the first researchers’ codes and or categories against a sample of the data. Differences were planned to be resolved by discussion.

## Ethical considerations

The study was approved by the Health Research Authority and Health and Care Research Wales (IRAS ref. 313346) and was not classed as research requiring NHS ethical approval within Health Research Authority guidance (HRA, 2019). Organisational (Governance) approval was received from the Medical and Clinical Services Directorate of the Welsh Ambulance Services University NHS Trust. Respondents were healthy adult volunteers and consent to participate was assumed by completion of the questionnaire. The data set supplied to the research team by YouGov was anonymous.

## Findings

The characteristics of this sample are presented in table 1 and 2.

**Table 1:** 2022 survey characteristics.

| **2022 Cohort’s characteristics** | **Overall Cohort** |
|---|---|
| **Total sample** | n= 1,010 |
| **Gender n (%)** |  |
| Female | 529 (52.4%) |
| Male | 481 (47.6%) |
| **Age** | 18-24: 81 (8.%)<br>25-34: 139 (13.8%)<br>35-44:179 (17.7%)<br>45-54: 148 (14.7%)<br>55+: 463 (45.8%) |
| **Social Grade n (%)** |  |
| ABC1 | 530 (52.5%) |
| C2DE | 380 (47.5%) |
| **Welsh region n (%)** |  |
| Mid and West Wales | 193 (19.1%) |
| North Wales | 209 (20.7%) |
| Cardiff | 133 (13.2%) |
| South Wales Central | 69 (6.8%) |
| South Wales East | 235 (23.3%) |
| South Wales West | 171 (16.9%) |
| **Working Status n (%)** |  |
| No | 524 (51.9%) |
| All workers (NET) | 486 (48.1%) |

**Table 2:**
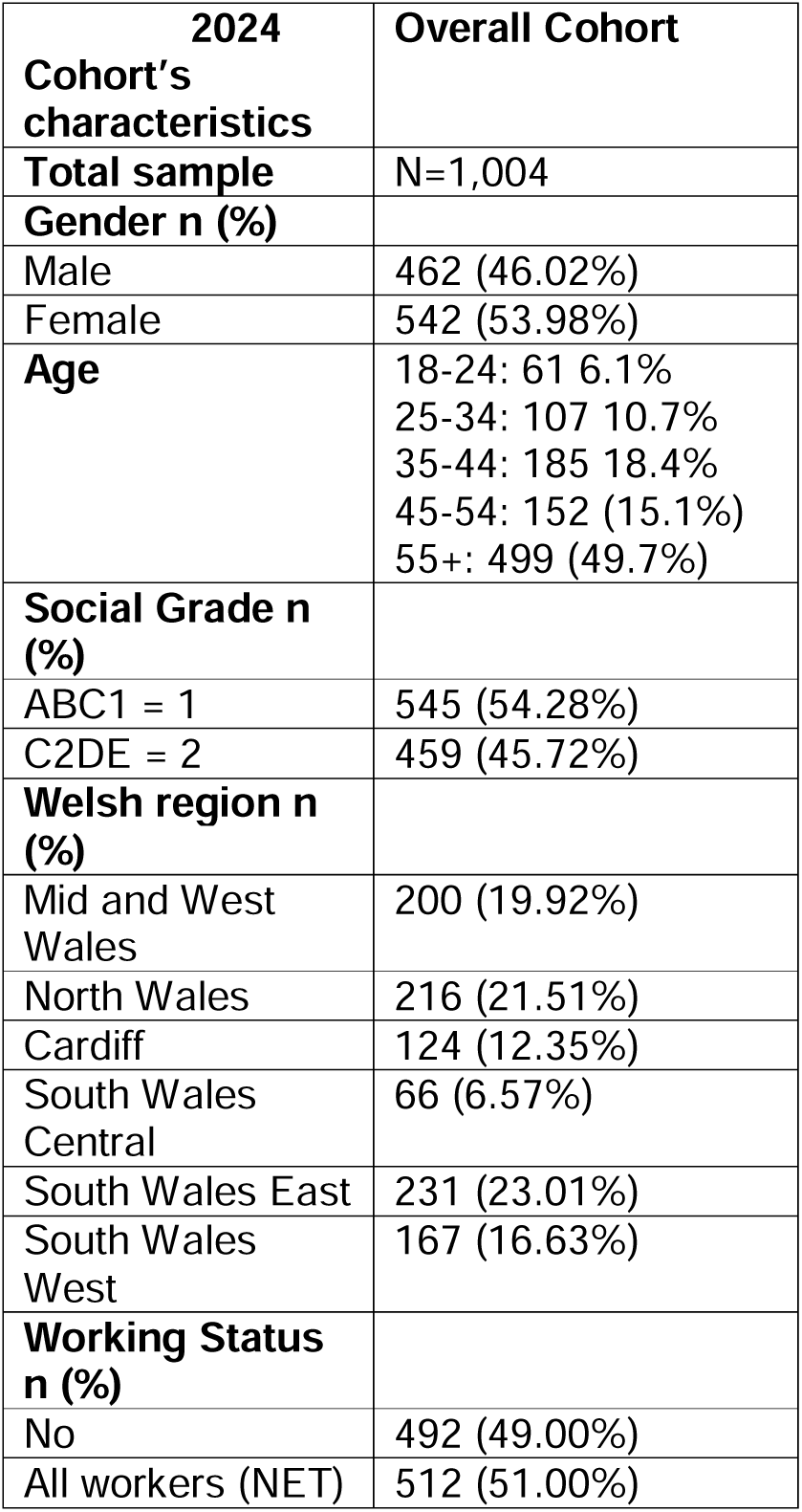
2024 survey characteristics.

| **2024 Cohort’s characteristics** | **Overall Cohort** |
|---|---|
| **Total sample** | N=1,004 |
| **Gender n (%)** |  |
| Male | 462 (46.02%) |
| Female | 542 (53.98%) |
| **Age** | 18-24: 61 6.1%<br>25-34: 107 10.7%<br>35-44: 185 18.4%<br>45-54: 152 (15.1%)<br>55+: 499 (49.7%) |
| **Social Grade n (%)** |  |
| ABC1 = 1 | 545 (54.28%) |
| C2DE = 2 | 459 (45.72%) |
| **Welsh region n (%)** |  |
| Mid and West Wales | 200 (19.92%) |
| North Wales | 216 (21.51%) |
| Cardiff | 124 (12.35%) |
| South Wales Central | 66 (6.57%) |
| South Wales East | 231 (23.01%) |
| South Wales West | 167 (16.63%) |
| **Working Status n (%)** |  |
| No | 492 (49.00%) |
| All workers (NET) | 512 (51.00%) |

Two thousand and fourteen free text narrative responses were analysed, and the following conceptual themes emerged:

### Multimodal, cumulative and confabulated knowing

Whilst respondents reported being aware of V&A communications, almost all could not recall actual campaigns, or where they received the message from. Only one of the 1010 respondents in the 2022 survey correctly remembered the WAST campaign as “*Work with us not against us*”. Other suggestions however included “*hashtag be kind*” and “*Respect and protect*”. Respondents appeared to confabulate and recall information that did not fit with actual EMS media campaigns:

*“I just remember seeing adverts and posters in hospitals, often depicting an EMS worker with bruises and the message that violence and aggression towards these workers will not be tolerated”*

We are not aware of any UK EMS media campaigns that included EMS workers with bruises, but this may have been the case for in-hospital campaigns. Respondents referred to campaigns such as:

*“Hashtag be kind”,* “*Respect and Protect”* And *“Don’t choose to abuse campaign”*

We cannot find evidence of these being actual campaigns and may be imagined or confabulations, where there may be no intent to deceive, but rather, distortions of information through memory error or misinterpretations. Respondents also generalised their responses to include V&A on staff across other organisations and occupational groups. They thought they had heard such information through embedded messages such as in phone line messages:

*“I don’t know the name of the campaign but I’ve been reminded of seeing adverts on TV in the past about showing respect to food workers”*

And

“*It came in line with many companies who use a menu system on their phones requesting that staff be treated with respect and courteous manners otherwise the call would be terminated and the call handler had the right to do this*”

Respondents did however overwhelmingly report encountering campaigns highlighting V&A directed towards EMS staff across many forms of media including television, newspapers, social media, and posters. This included news programs, documentaries, the television drama casualty, social media such as Twitter [now X] and Facebook, and in settings such as medical centres, hospitals, buses, and trains:

*“Ddim yn cofio enw’r ymgyrchoedd. Wedi gweld deunycentersddiau ar y cyfryngau cymdeithasol ac ar y teledu. Clywed gan ffrindiau a theulu hefyd”*

*[Can’t remember the name of the campaigns. Have seen materials on social media and on television. Heard from friends and family too]*

One respondent also appeared to accurately reflect the message and means of communicating the #*Workwithusnotagainstus* campaign (WAST 2026) and stated:

*“I’ve seen a poster on an ambulance car with the image of someone threatening a worker but can’t remember the quote. I’ve also seen it outside a hospital”*

The #*Workwithusnotagainstus* (WAST 2026) campaign did indeed use images on ambulance cars to promote this message along with posters in hospitals. Key messages within campaigns were also reported by respondents such as the demands of the job they do:

*“an advert explaining what these workers sadly have to go through while just doing their job”*

And the need to show respect for EMS staff:

*“I have seen adverts asking people to respect emergency workers”*

And, the potential punishment those who abuse EMS staff may receive:

*“Main focus is to make people aware they can be punished for abuse against EMS staff”*

Respondents overwhelmingly viewed V&A towards EMS staff as socially unacceptable and shared further reflections and personal thoughts towards this:

*“I don’t remember any details but have seen publicity on TV and online urging respect. I remember commenting at the time that it was awful that it was necessary to run such a campaign”*

And

“*It’s sad that such a campaign should have to be run*”

The 2024 survey may reflect a more encouraging and gradual effect of media campaigns as the Wales media campaign #*Workwithusnotagainstus* (WAST 2026) was mentioned four times and the England media campaign ‘*Work without fear*’ (AACE 2026) was mentioned five times. Attributing these responses to individual campaigns however is difficult, and many of the messages may reinforce already-held pro-social attitudes to this issue. Many also reported personal experiences of being assaulted as a health worker or by talking to friends and families of those who had been assaulted themselves or witnessed it:

*“My son in law was a full-time EMS and my daughter a staff nurse in A&E so I hear about incidents”*

Zero tolerance featured in over thirty responses:

“*Zero tolerance to people not showing respect to all emergency workers*

And

“*Zero tolerance towards verbal and physical attacks on emergency workers*.

### Dual discourse of V&A directed towards EMS staff

Respondents’ responses to the survey reflected simultaneously two conflicting and contrasting narratives involving reprehension and the social unacceptability of V&A directed towards EMS staff while also acknowledging causes such as frustrations over ambulance delayed response, the behaviour of EMS staff, medical conditions such as dementia, mental health, pain and alcohol. Respondents conveyed a sense of reprehension and overwhelmingly reported disgust and revulsion at V&A directed towards EMS staff and perpetrators were referred to in pejorative terms such as “*scum*” and “*idiots”*:

*“What I had seen on the news find it disgusting that scum like this get away with it time after time and they come out with the mental health card, people doing their jobs and helping the public and have to put up with these idiots”*

The reprehension seemed to extend beyond the actual V&A incident itself and the impact on the EMS staff to reflect wider issues such as the role EMS staff play within the community in saving lives and the impact it could have on individuals:

*“Treat the staff with respect. They are probably trying to save a life. It could be a member of your family next time”*

The reprehension of V&A towards EMS staff and its perpetrators contrasted with the high regard for EMS workers. Responses cited the increased dialogue within the public domain during the COVID-19 pandemic regarding the role EMS staff played and the hope for greater respect for what they do.

No respondents correctly recalled or cited the Assaults on Emergency Workers (Offences) Act (2018). There was however some awareness and support for legislation aimed at protecting EMS staff as it was mentioned nine times:

*“law is in place to protect them, although after it happens is always too late”*

And

*“Antagonism towards these people who are trying to help is disgusting, criminal and inhuman. Either help or, at least, keep clear!”*

And

*“those who are prosecuted face jail”*

While such changes in legislation have been included in communication campaigns, two respondents cited the high-profile killing of Police Constable Harper whilst on duty:

“*After the death of the police officer Andrew Harper*”

And

*“NHS and Harper’s law campaign”*

Respondents offered potential suggestions or situational triggers for increases of V&A towards EMS staff which reflects the context of EMS care and is rarely included in public discourse and communication campaigns. This included frustrations of accessing care and waiting for EMS to arrive:

*“When people got to wait a long time for EMS to turn up over 12 hours”*

And

*“8 weeks ago I spent 14hrs in an A & E after having a bad fall. Member of the public phoned for an ambulance but was told there were no ambulances (all queueing at the hospital) for 3hrs - I was taken to hospital by car. People waiting there sooo frustrated”*

Resorting to V&A in such circumstances is clearly unacceptable and would not be tolerated in other areas of society. Respondents did however point out that this was not a one-sided discussion; that V&A may be subjective and EMS staff could also have a role, which is rarely reflected in the public discourse and media campaigns:

*“We have been at the hands of rude, cruel EMS staff both in home and in Hospital. Unfortunately, we had to voice our unhappiness which could be misconstrued these days as “aggression”*.

And:

*“many ill people are afraid and may act aggressively and those suffering from Dementia can become aggressive and physical when disorientated. It’s not nice but it happens”*

Whilst the public overwhelmingly reported disgust at V&A towards EMS staff there appeared to be recognition and understanding of the complex context within which V&A occurs which belies a kind of pleuritic view of heinous perpetrators of V&A on heroic EMS staff. Respondents reflected the real-world context of EMS work and suggested that while V&A directed towards EMS staff may be reprehensible and not acceptable it may be expected and somewhat predictable:

*“Ambulance workers deal direct with members of the public in the worse conditions frequently and when people are drunk or on drugs they can’t distinguish from someone trying to beat them or help them”*

And:

*“Violence is not acceptable however the very nature of the role means employees may sometimes be dealing with people who are mentally ill and unable to control or understand their violent action at the time for what it is for example if they are having an episode, delusional or hallucinatory. If they have previously suffered an abuse of power from an authority figure especially a medical professional they may react instinctively and take time for trust to be established”*

And:

*“ If they have previously suffered an abuse of power from an authority figure especially a medical professional they may react instinctively and take time for trust to be established. Aggression is sometimes acceptable for the same reasons and because it is only natural that when people are scared, for example when they are hurt, they may react with bad language or shouting which may or may not be directed at the employee trying to help them. While this may be scary or upsetting for the employee, this is to be expected if you choose to enter this type of profession”*

Respondents to the survey appeared to reflect the reciprocal duty society and employers must protect EMS staff in turn for providing this vital service to the community. This responsibility extended to mitigate the personal, physical, and psychosocial harms of such V&A:

*“employees at all levels should be trained specifically in how to cope with these scenarios through realistic role play, trained in coping with any personal mental effects, paid higher salaries to reflect the higher stress/unpleasant aspect of the job, provided with free optional discreet protective padding for beneath their uniforms and complimentary unlimited access to psychological therapies throughout and after their career ends”*

Figure 1. presents a truncated example of coding:

**Figure 1:**
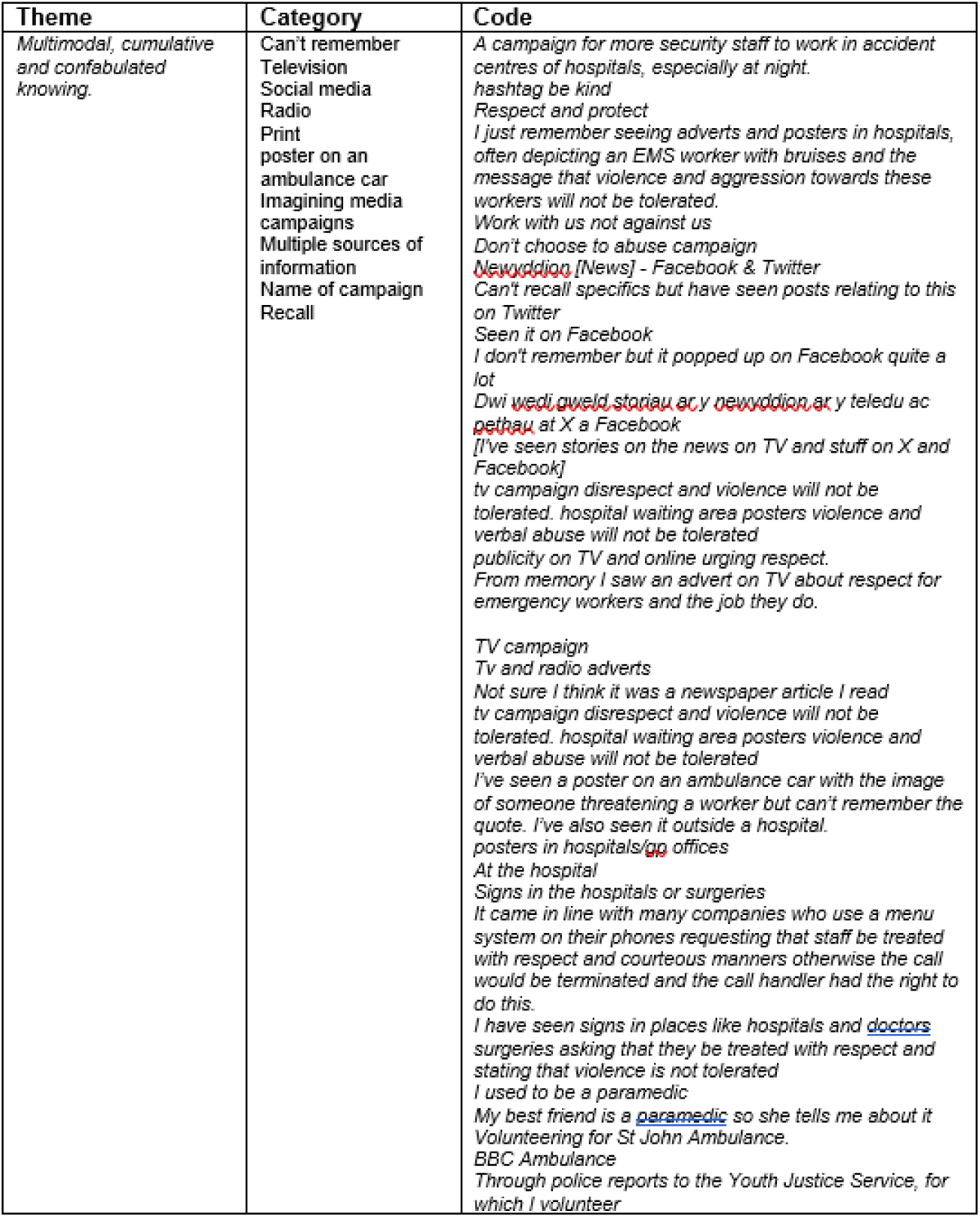
Truncated example of coding:

## Discussion

Respondents’ reported unacceptability and revulsion at V&A directed towards EMS staff, which is not surprising and is supported by the quantitative findings from this survey and our qualitative research exploring the experiences of UK EMS staff (Rees et al 2025, Rees et al 2026). Respondents also appear to support prosecution and harsh criminal sentences in such circumstances, and whilst none cited the Assaults on Emergency Workers (Offences) Act (2018), there was some awareness and support for harsher sentences for assaulting emergency workers. Two respondents referred to Harper’s Law, which is UK legislation mandating life imprisonment for anyone convicted of the manslaughter of an emergency worker who is killed while on duty (Police, Crime, Sentencing and Courts Act 2022).

Few would dispute that the introduction and enacting of such specific legislation for these criminal offences is fitting punishment and an understandable societal response reflective of the level of moral outrage. Ambulance services and the NHS have stepped up communication campaigns in recent years, but it is unclear what effect these may have over and above already held knowledge and attitudes of revulsion and disgust?

Most people in our survey could not recall actual media campaigns aimed at reducing V&A on EMS staff. In the 2022 survey only one referred to the major campaign in Wales #*Workwithusnotagainstus* (WAST 2026) which had recently been promoted extensively to protect EMS staff from V&A. The 2024 survey may reflect a more encouraging and gradual effect of media campaigns as #*Workwithusnotagainstus* (WAST 2026) was mentioned four times and the England media campaign *Work Without Fear* (AACE 2026) was mentioned five times (despite the survey including only a Wales population). Despite this limited awareness, responses reflect the messages included in these campaigns, alongside a combination of understanding acquired through many forms of personal experiences, social media, television and posters. Such transmedia knowing involves telling stories that unfold and extend across multiple platforms and has the potential to go deeper as they gain different points of view related to the story world (Jenkins et al 2006).

Media effects include theories to explain how media influences attitudes and perceptions and represents one of the core ideas of communication research (Neuman & Guggenheim 2011). Historically, media effects were characterized by either “*significant*” or “*minimal*” (Bryant &Thompson 2002, McQuail 2010). The boundaries of such effects are defined by emerging media technologies, culture, types of methods, perspectives, and ideologies used. Significant media effect proposed in the1920s often incorporated “*magic bullet*” metaphors, as if messengers shoot messages and effects into receivers; media were credited with immense power in shaping opinion, attitudes, and behaviour. Harold Lasswell (1927) used arguments of Freudian psychology, pragmatism, and political science in suggesting that society and individuals could be effectively controlled by a minority through propaganda. Joseph Klapper (1960) however argued that the effects of mass communication merely reinforced predispositions and do not directly influence them.

More recently minimal models of media effects place emphasis on constructivist models (McQuail 2010), suggesting that much of what the media do involves the construction of reality, with the public deciding whether to adopt the media’s world view or not. Constructivism is a theory of meaning-making that enables people to create their own new understanding based on an interplay between what they already know and believe and the concepts and knowledge with which they interact (Resnick& Glaser, 2016).

Members of our team have applied constructivist philosophical approaches in EMS research in areas ranging from mental health to the COVID 19 pandemic (Rees et al 2016, 2017, 2018, 2021 a, 2021 b, 2021c, 2022, 2026). Powerful concepts support the notion of constructed knowledge in relation to EMS are reflected in this study and our previous work. We therefore argue that as with other areas of EMS care, and social interaction, people use a wider range of personal and prior knowledge or experience of EMS work and V&A to construct new understandings, modified through information and experiences, which is an active rather than passive process.

Respondents in our survey displayed some mental engagement with the topic area, whilst these are limited to short narrative responses, rather than being passive respondents’ and recipients of information, they shared prior knowledge and synthesised this with new knowledge with their thoughts and experiences into new realities. The sophisticated insights into the dual discourses reflected such meaning-making occurring both through personal reflection and social interaction. We recognise however that how the media frames narrative related to health professionals matters and can influence the public’s perceptions of those people, services and how they discuss public events. Multiple campaigns over many years, personal knowledge, latent metaphors such as zero tolerance may also represent a cumulative effect where such heuristics are sometimes confabulated to include non-existent campaigns.

While these responses may reflect efforts to tackle this issue through communications campaign such as #withusnotagainstus (WAST 2026), given the dramatic nature of the images and related warnings of harsh sentences that are employed, these campaigns clearly capture audiences’ attention and responses. Combining fear and disgust are viewed as particularly effective in public health campaigns in terms of drawing attention to the health threat (Wu and Morales 2012).

The linking of fear and risk with emotion is often a central strategy of health promotion, and V&A directed towards EMS staff is indeed emotive and of high public interest. Lupton (2013) described such an ‘emotion-risk assemblage’ or the configuration of human and non-human actors that combine to give meaning to both risk and emotion. Our study reflects such an emotion-risk assemblage where V&A directed towards EMS staff yields powerful and emotional public responses of revulsion and disgust due to risk of harm to EMS staff, who are held in high public regard and characterised as heroes (Rees et al 2022). Conversely V&A directed towards EMS staff results in potential loss of their vital services to the public which are deemed to be lifesaving, and therefore presents personal risk to themselves, their families and communities.

We have previously reported that it is such personal risk of illness or injury through V&A, whilst continuing to care for patients sustains the characterisation of EMS staff as heroes (Campbell1968, Rees et al 2022). EMS staff however appear ambivalent to such heroism narratives, due to politicisation, and objectification in the media and society (Rees et al 2022, Tangherlini 2000). Rather, Paramedics have been reported to present themselves as anti-heroes, quick for a sardonic quip (Tangherlini 2000). Cox (2020) reflected such discourses in other health workers, arguing that heroism narratives stifles meaningful discussion about the limits to healthcare professionals’ duties and treatments, fails to acknowledge the importance of reciprocity, and can have negative psychological effects on workers themselves through its implication that all healthcare workers have to be heroic.

Media campaigns and reports employing disgust may be amplifying such emotion risk assemblages, in which the already emotionally-resonant meanings of risk are bestowed with additional power through their association with revulsion. The logic underpinning the use of such images to evoke intense emotional responses from target audiences is that members of these audiences are apathetic or resistant to the health messages public health authorities are attempting to convey to them (Crawshaw 2012; Gagnon, Jacob and Holmes 2010; Lupton 1995, 2013a, 2014). Employing the pedagogy of disgust in such messages may combine facts and figures to bolster their authority, but they are essentially attempting to elicit a negative affective response in a very overt manner.

Health education materials and the media are replete with messages referred to as ‘shock tactics’ or ‘distressing’, ‘disturbing’ or ‘graphic imagery’ to provoke behaviour change in target audiences (Lupton 2014). We have previously reported how people who use drugs or alcohol and assault EMS staff often feature in media reports as ‘drug addict’, in a ‘drug-addled stupor’ and aligned with ‘thugs, perverts, drug dealers and other criminals’ (Rees et al 2022). Whilst our study revealed less pejorative aspects in ambulance services communications, with terms such as be kind and show respect, these were in the minority of more negative responses to our survey such as zero tolerance which featured in around thirty responses.

We are not aware of media campaigns using the term Zero Tolerance recently, but this was the stance of V&A directed towards NHS staff communicated throughout the 2000’s (Whittington et al 2002). No explanation was provided of what zero tolerance meant, along with lack of underpinning policy and legislation at the time to tackle V&A, and evidence, suggests the approach was ineffective (Whittington et al 2002, Rees et al 2025, Weeks et al 2002). This reflects findings of other research on communication campaigns related to violent incidents at the time that similarly did not achieve their goal of reducing the problem (Cárdaba 2016). Indeed, messages that contain prohibitions or seek to change behaviour through fear appeals tend to be ineffective or even counter-productive for reducing violent attitudes and aggressive behaviour (Cárdaba et al 2016, Bushman & Stack 1996, Roskos-Ewoldsen 2004).

The effectiveness or otherwise of efforts to reduce V&A directed towards EMS staff through media campaigns and legislation requires careful ethical deliberation which is beyond the scope of this paper. However, the two dominant normative theories of consequentialism and retributivism offer contrasting yet complementary lenses through which this can be examined. Consequentialism (Beccaria 1764 and Bentham1789) asserts that punishment is justified by its outcomes, specifically its capacity to reduce future harm through deterrence, rehabilitation, or societal protection. Retributivism, in contrast, grounded in Kantian moral philosophy and further theoretical developments by Moore (1987), Duff (2001) and others, posits that punishment is morally justified when it gives offenders what they deserve, based on the severity of their wrongdoing. This theory emphasizes the intrinsic moral obligation to hold perpetrators accountable, independent of the consequences, reflecting the principle of justice as fairness (Landa 2009, Stern 2024).

Kiettikunwong et al (2025) contends that the moral architecture of the UK Assaults on Emergency Workers (Offences) Act (2018) is overtly retributivist, where for instance, government ministers emphasize that an attack on an emergency service provider is considered an attack on the social order itself warranting punishment more severe than an ordinary assault on a private citizen (Crown Prosecution Service 2025; Weeks and Broughton 2023). A strength of such a legal response suggests Kiettikunwong et al (2025) is its clarity and communicative function, sending out an unambiguous public message that abuse or violence toward healthcare workers is unacceptable and will be met with the full force of the law. This clarity suggests Kiettikunwong et al (2025) can empower NHS institutions to enforce a “zero tolerance” culture and provision of notices warning that aggression toward staff may lead to arrest and prosecution.

The Association of Ambulance Chief Executives (AACE 2026) and UK ambulance services have refreshed efforts to reduce V&A directed towards EMS staff which reflect a hybrid orientation, highlighting legal responses to healthcare violence, that combine punitive sanctions with preventive and supportive measures such as mandatory reporting systems, support services for affected workers, training, body worn video and workplace violence prevention programmes. While AACE calls for further interventions, few evidence based interventions exist. AACE also report the highest recorded levels of V&A reaching 22,500 a year (2024 – 2025 period) for the first time ever. As with the zero tolerance approaches in the 2000’s such figures may therefore reflect the limitations of the legislation and current approaches to stemming the tide of V&A directed towards EMS staff?

Much progress has undoubtedly been made from a retributivist perspective in its moral condemnation and public symbolism. Consequentialist objectives however call for evidence of deterrence of future harm through such enhanced sentencing and visible enforcement (Weeks and Broughton 2023; Parpworth 2022). Under this view, laws and approaches to healthcare violence should be designed to prevent future incidents, reduce recidivism, and promote institutional safety (Wood 2010; Pilcher et al. 2024).

Respondents in our survey recognised the dual discourses at play with this issue, and thought intoxication with alcohol (92.4%), drugs (90.5%) and altered mental status following illness and/or injury (84.3%) would likely contribute to V&A towards EMS staff. Interestingly drugs and alcohol are identified as aggravating factors (justifying harsher sentencing) in the Assaults on Emergency Workers (Offences) Act (2018), whereas mental health problems are seen as mitigating factors (warranting more lenient sentencing). Despite this, respondents’ thought the act would unlikely deter perpetrators who were intoxicated with drugs (75.2%), alcohol (75.2%), with altered mental status following illness and/or injury (75.6%) or other member of the public (42.4%) (Rees et al 2025). Despite this, public and policy discourses on V&A directed towards EMS staff rarely feature the voice of the perpetrator who may also be the patient. A power imbalance may therefore exist in terms of who establishes the narrative, and how is this is systematized matters. Media reporting of people who use drugs or alcohol and assault EMS staff use words such as ‘*drug addict*’ ‘*thugs, perverts, drug dealers and other criminals*’ (Rees et al 2022), but we argue that such binary characterisation and valorisation of heroic EMS staff versus de-valorisation of heinous criminals is a gross oversimplification and may be unhelpful.

Viking et al (2020) explored V&A in the ambulance service from the offender’s perspective and conducted a qualitative study using trial transcripts from cases tried in court. Four themes were found: (I) the offender was misunderstood, which included communication problems and other misunderstandings; (II) the offender was disrespected, which described perceived unprofessional behaviour and unpleasant or painful treatment by ambulance personnel; (III) the offender was vulnerable, which described the state of the offender (i.e., being under the influence of alcohol or drugs); and (IV) the offender had unmet expectations, which included perceived unreasonable waiting time and conflicting expectations of ambulance care.

Spelten et al (2020) conducted focus groups to identify and discuss the perceptions of Emergency Department nurses, about perpetrators of V&A. Respondents’ acknowledged that V&A had become an intrinsic part of their job and identified the following six overlapping groups of perpetrators: 1: Violence or aggressive behaviour that cannot be explained by an underlying health issue 2: Violence that is related to underlying mental health issues. 3: Violence is related to underlying physical health issues 4: Violence that is related to addiction and substance abuse 5: Violence that is related to a complexity of issues. 6: Violence that is related to repeat visitors/offenders.

Nurses in Spelten et al (2020) recognised that it was not within their power to solve this issue given the societal component, and identified several organisational/societal factors they thought contributed to the incidence and nature of violent incidents, including: waiting times as a source of frustration and irritation, cultural barriers, particularly in relation to a predominantly female workforce, social media, the instant posting and the public ‘naming and shaming’ was considered a major issue and made the nurses feel very vulnerable. Spelten et al (2020) called for the attention to shift from equipping workers with tools to manage violence to the perpetrator and the development of interventions to reduce violence from targeted perpetrator groups.

Evans et al (2023) recently explored experiences and views of people who frequently call emergency ambulance services. This population are often vulnerable, more likely to experience mental health challenges including self-harming behaviour, live with chronic conditions or misuse drugs and alcohol. In this study respondents said they felt criminalised by the way they were treated by emergency services. Four of the six were attended by police officers when calling for an emergency ambulance. One said she was arrested for violent behaviour linked to a psychotic and self-harming episode. One respondent also said he was threatened by the ambulance staff for refusing treatment during a mental health crisis because of his fear of hospitals. “*It’s like they threaten me for refusing to go there, isn’t it? They start mentioning things like, ‘Oh, you won’t get benefit anymore because you’re not taking our advice and our services’…I’m petrified of hospitals* (Watkins et al 2025, Evans et al 2023). Evans et al (2023) suggested that police involvement was a normalised element of their experiences. This reflects previous findings from interviews with paramedics we conducted that revealed similar involvement of police and surreptitious strategies related to care and detention of people who self harm and ’ways and means’ that have evolved to overcome complexities of care (Rees et al 2017).

The responses to the survey reflect such a dual discourse, where a minority of respondents suggested that EMS organisations and their staff may well also have a role in V&A directed towards them. This represents a contrasting and more complex narrative, involving wider systemic frustrations we have reported previously, which involved ambulance delayed response, the behaviour of EMS staff, medical conditions such as dementia, mental health, pain and alcohol (Rees et al 2025). These issues remain largely silent in the public discourses and rarely feature in media campaigns, which also lack an empirical underpinning evidence base. We have previously reported the socio-political dimension to this issue and how newspaper coverage can be reductionist with a strong tendency to either attribute blame to the patient behaving aggressively by NHS management and being seen to be acting or so that politicians may win more votes by being ‘tough on crime’ (Eburn et al Hoyle et al Rees et al 2025, 2026).

In our interviews with EMS staff the following categories emerged which lay bare such dual discourse and provide stark contrast to public narratives: *Rusted, busted and inevitability of V&A in EMS environment, Tolerable or intolerable V&A in EMS, Gendered V&A and genderisation in EMS, Modifiable factors and harm reduction of V&A in EMS, Professional, ethical and clinical judgments of V&A in EMS and Sociocultural and system frustrations of V&A in EMS.* The basic social process that emerged was one of: *Systemic frustration and gender-based V&A in EMS*.

Respondents in our survey (Rees et al 2025) also offered potential suggestions for the rises in V&A directed towards EMS staff which included delayed response, being in pain, scared and confused, limited mental capacity due to drugs, alcohol, medical and mental health problems which reflects the complexity of V&A directed towards EMS staff previously reported by ourselves and others.

The Royal College of Emergency Medicine recently released *Safe to Care Violence and Aggression against Emergency Department Staff* (RCEM 2026). This campaign included a UK-wide survey of emergency department staff which found rising levels of V&A occurring daily in emergency departments across the UK. The campaign included many of the stark statistics and messages that V&A is unacceptable. This campaign highlights that individuals must always be held accountable for criminal behaviour, reflecting retributionist principles discussed earlier. They also however recognise that some incidents involve patients experiencing delirium or a mental health crisis which further reflects the complexity and wider system issues we have previously reported which rarely features in such campaigns. RCEM (2026) acknowledges the dual discourses and how:

*“Violence and aggression are not simply isolated incidents caused by patients, but rather, are increasingly associated with: sustained overcrowding, staff shortages and deteriorating emergency care environments. These conditions increase frustration, reduce privacy, place staff under greater pressure and make incidents more likely”*.

RCEM (2026) therefore reflects a more consequentialist approach to prevention, highlighting the reciprocal responsibilities for governments across the UK and NHS organisations to reduce the conditions that make violence more likely, ensuring that the nature of emergency medicine and the risk of V&A is recognised, with everyone possible done to keep staff safe. RCEM (2026) calls for a range of actions to prevent V&A before it occurs by tackling overcrowding, poor environments and long waits. Introduce national standards for security in ED’s, provide support to victims of V&A, ensure perpetrators face justice – taking a cross-system approach across the NHS/HSCNI, police services and the crown prosecution service and devolved equivalents.

Such discourse rarely features in media reports, however in a recent article in the Guardian newspaper Professor Nicola Ranger (general secretary of the Royal College of Nursing RCN), said:

*“while patients with severe mental health problems, dementia and delirium were responsible for many assaults on staff, there was also growing deliberate aggression from other patients due to long waits for care”*

And

*“The cause of their violence is the poor service that we are giving them and all their family members,”*

And

*“That can only be prevented if the service improves.”* (Batty & Bawden 2026)

## Conclusion

Respondents overwhelmingly reported the unacceptability of V&A directed towards EMS staff, which reflects public discourses replete with disgust and revulsion. We found limited awareness of media campaigns aimed at tackling V&A. Responses did however reflect messages included in campaigns but these insights were acquired through a multimodal and cumulative process reflective of constructivist theory of learning where people create understandings based on an interplay of social interaction, what they already know and believe, personal reflection and the concepts and knowledge with which they interact.

Respondents appear to support prosecution and harsh criminal sentences for perpetrators of V&A towards EMS staff and whilst none cited the Assaults on Emergency Workers (Offences) Act (2018), there was some awareness and support for its aims. It is reassuring therefore from a retributivist perspective that public policy may be aligned with societal and moral expectation. It is unclear however what media campaigns have added to this discourse? If their purpose is to communicate the unacceptability of V&A towards EMS staff, messages of zero tolerance and harsh sentences for perpetrators, there appears to be little space to add to already held public attitudes and insights. We also found limited evidence of the preventative capacity of current approaches, which is consistent with our previous findings and wider body of knowledge. We therefore call for future research and policy to focus on more complex and considered solutions to tackling these issues. Such approaches should recognise the dual discourse revealed in our study and follow more consequentialist objectives to preventing future incidents, reducing recidivism, and promoting safety as opposed to the highly visible media headlines.

## Data Availability

All data produced in the present study are available upon reasonable request to the authors

